# A multi-method study to identify and validate alcohol diagnostics for injury-related trauma in South Africa: a study protocol

**DOI:** 10.1101/2022.12.19.22283649

**Authors:** Petal Petersen Williams, Megan R. Prinsloo, Margaret M. Peden, Ian Neethling, Shibe Mhlongo, Sithombo Maqungo, Charles Parry, Richard Matzopoulos

## Abstract

**Introduction:** The burden of alcohol use among trauma patients and the relative injury risks is not routinely measured in South Africa (SA). Given the prominent burden of alcohol on hospital trauma departments, SA needs practical, cost-effective and accurate alcohol diagnostic tools for testing, surveillance and clinical management of trauma patients. This study aims to validate alcohol diagnostics for injury-related trauma and assess its utility for improving national health practice and policy.

**Methods and Analysis:** The Alcohol Diagnostic Validation for Injury-Related Trauma (AVIRT) study will use mixed methods across three work packages. Focus Group Discussions will be conducted with key stakeholders across four areas of expertise (clinical, academic, policy and operational) to determine the type of alcohol information that will be useful for different stakeholders in the injury prevention and healthcare sectors. We will then conduct a validation study of alcohol diagnostic tools (clinical assessment, breath analysis, finger-prick blood) against enzyme immunoassay blood concentration analysis in a tertiary hospital trauma setting. Finally, selected alcohol diagnostic tools will be tested in a district hospital setting alongside community-based participatory research on the utility of the selected tools.

**Ethics and dissemination:** The study was approved by the Research Ethics Committee of the South African Medical Research Council and the Western Cape Health Department. Findings will be disseminated to inform strategies to introduce routine, cost-effective alcohol diagnostics optimally in a high trauma setting by ensuring accuracy, real world feasibility and institutional support.

**Strengths and limitations of this study:**

- The study will provide an understanding of the impact of alcohol and its association with injuries, highlight the burden alcohol-related injuries impose on health workers, inform policies to mitigate alcohol harm, and recommend methods to scale-up alcohol screening and detection in trauma patients nationally.
- The study setting will complement the limited evidence base on alcohol consumption in LMICs.
- Expert stakeholders included in focus group discussions will be restricted to known contacts within the field.
- Validity testing of the alcohol diagnostics will use enzyme immune assay instead of the gold standard gas chromatography method, which was too expensive.
- Results will be generalisable to patients treated at public-sector district and tertiary-level facilities, that treat a higher proportion of moderate to severe injuries.

## INTRODUCTION

Alcohol indicators obtained from patients seen in emergency departments or admitted to hospital are one of the key and most cost-effective data sources to estimating the impact of alcohol on communities and health [1] to quantify problem drinking, alcohol-impaired driving, trauma readmissions and premature death [2]. This assists in identifying high risk groups that should be targeted for prevention. Effective monitoring of alcohol-related morbidity and mortality requires the collection of alcohol-related indicators in which regular reports on the key predefined indicators are submitted by hospitals, primary health-care units or emergency services [3, 4].

The contribution of alcohol to the global burden of disease is undisputed. In addition to risks such as non-communicable diseases, infectious diseases and mental health problems as a result of hazardous and harmful alcohol use [3], the trauma burden of intentional and unintentional risk of injury from alcohol is a major public health concern. Studies from sub-Saharan Africa have highlighted the impact of alcohol on injury and violence [5-8] and concerns over alcohol consumption and alcohol-attributable burden of disease. The lack of attention alcohol-related harm receives from policymakers has been raised [9, 10], with calls for stronger and more effective alcohol control measures.

In South Africa (SA), approximately a third (31.2%) of alcohol-attributable deaths in 2012 occurred as a result of injuries; while 15.9% and 12.8% of alcohol-attributable disability adjusted life years (DALYs) were caused by road traffic injuries and interpersonal violence respectively [11]. The adult per capita consumption of alcohol in SA is extremely high (64.6 grams of absolute alcohol (AA) per drinker per day). Almost 6 out of 10 South African drinkers over the age of 15 years are reported to engage in heavy episodic drinking (HED) [3], which is strongly associated with increased injury risk.

COVID-19, and the related alcohol-bans in SA, has brought the impact of alcohol on trauma presenting to health facilities into sharp focus in the country [12]. Moultrie et al. (2021) [13] and Barron et al (2022) [14] demonstrated how a total ban on alcohol during the current COVID-19 pandemic resulted in significantly fewer injury deaths. However, the absence of routine and reliable alcohol-related injury surveillance data have been identified as a critical gap and the government has had to rely on the South African Medical Research Council’s (SAMRC’s) rapid mortality reporting [15] of all injury-related deaths and ad hoc surveillance studies [13, 16] to demonstrate the association between the availability of alcohol and alcohol harm. This gap has substantially hindered the ability to provide regular information on changes in the pattern of alcohol related injuries and limits the ability to measure the impact of policy changes, implementation and enforcement.

As the country transitions from the COVID-19 crisis response, it is likely that there will be further pressure on the government – from civil society and health and social agencies – to implement more sustained intervention strategies to reduce harmful drinking and to monitor the impact of any interventions on the alcohol-related injury burden. This has highlighted the absence of practical, cost-effective and accurate alcohol diagnostic tools in the South African trauma setting. Accurate measurement would improve surveillance and influence the clinical management of trauma, inform and improve government policies to address heavy drinking and assess the impact of alcohol policy reform. The proposed study thus aims to determine the type of information that will be useful for stakeholders in the trauma care and injury prevention sectors; to validate the efficacy of a selection of alcohol diagnostic tools; and to explore their feasibility for wider provincial or national implementation as a routine source of information on the alcohol-relatedness of injuries.

## METHODS AND ANALYSIS

### Study design and setting

We will use a mixed methods participatory approach across three work packages (WPs) to validate alcohol diagnostics for injury-related trauma and assess its utility for improving national health practice and policy (Figure 1). Outcomes from this research will inform health practice, policy development and sustained intervention strategies. A validation study will be conducted in the trauma unit of Groote Schuur Hospital in Cape Town – a tertiary hospital. Further testing will be conducted in a district hospital in Mitchells Plain on the outskirts of Cape Town – approximately 28 km from the city centre. These two hospitals represent high injury caseloads, particularly for violence and road traffic injuries [17, 18].

**Figure 1:**
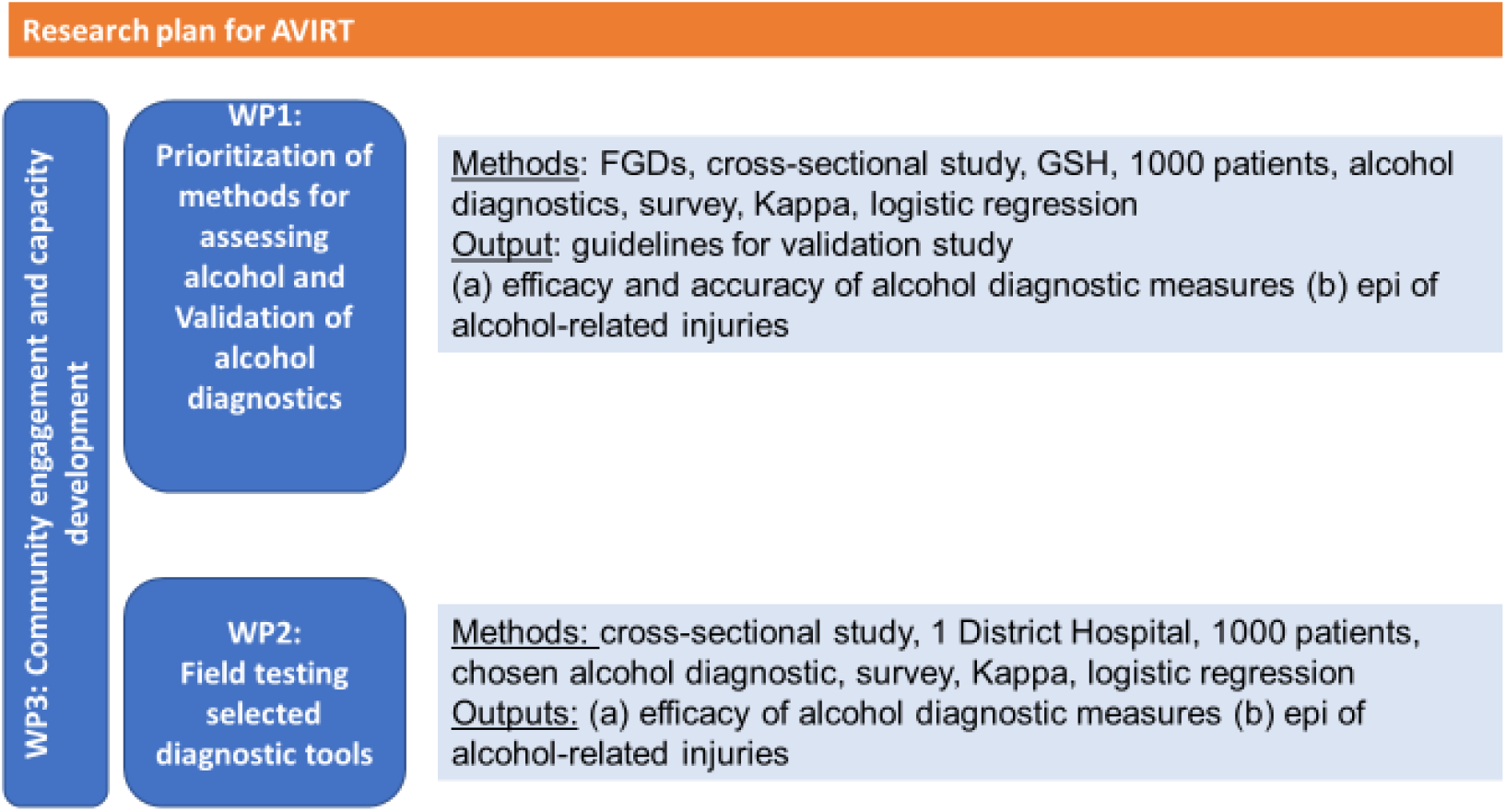
Diagrammatic illustration of the three studies and anticipated outputs. AVIRT = Alcohol Diagnostic Validation for Injury-Related Trauma, GSH = Groote Schuur Hospital

### Research plan

#### WP 1: Prioritization and validation of methods for assessing alcohol use

We will conduct Focus Group Discussions (FGDs) with various stakeholders to ascertain the current alcohol assessment practice in hospitals and the type of information that could assist in the acute management of injured patients and measure alcohol use for public health surveillance. This will be followed by a cross-sectional study to validate the efficacy of three diagnostic tools:

1. Clinical assessment, which is based on a clinician’s observation of apparent intoxication classifies the alcohol status of a patient into four categories as per ICD-10 coding (Y91.0 mild; Y91.1 moderate; Y91.2 severe; Y91.3 very severe); [19]
2. Breathalyzers provide an estimation of the ethanol content in the breath; [20] and
3. Surescreen Alcostick™ uses a finger-prick to provide a capillary blood reading in 1-2 minutes and is used to facilitate the clinical process, as well as the use of the ICD-10 Y91 coding [19].

These three index tests will be measured against venous blood testing for the presence of ethanol as the reference standard, for which we will use enzyme immunoassay [21] for testing of Blood Alcohol Concentration (BAC). The analysis will be conducted by a private pathology laboratory.

This WP will also describe the epidemiology of alcohol-related injuries among trauma cases presenting at the emergency unit of the selected trauma unit of a tertiary hospital in Cape Town through a prospective study. Patients presenting to the trauma unit for first time treatment of their injuries will be interviewed to record patient demographics, the injury intent and related mechanism and will be tested for alcohol, using the aforementioned alcohol diagnostic tools. Prior to the validation study, a small pilot study will be conducted over 2 weeks, to test the consent procedures, the study questionnaire platform, and the logistics surrounding the blood withdrawal and use of a courier to the contracted laboratory for centrifugation within the 2-hour limit to preserve the alcohol.

#### WP 2: Field testing selected diagnostic tools

Guided by findings from WP1, the alcohol diagnostic tools will be field tested in a district hospital in a large suburb on the outskirts of Cape Town. This WP aims to test the suitability of validated alcohol diagnostic methods for routine use in a hospital trauma setting on a day-to-day basis.

#### WP 3: Community engagement and capacity development

Engagement with clinicians, operational stakeholders (e.g. Non-governmental Organizations (NGOs) and emergency services) and individuals working in the policy arena are key to the success of any policy change. These stakeholders will be engaged through the FGDs at the commencement and again at the end to include community and patient representation. Three sets of activities for this phase will thus include:

- Step 1 - FGDs on the utility of the recommended methods and what would be required for implementation and the role of the measure in health care provision;
- Step 2 - workshops/webinars with stakeholders for input on findings and to convey recommendations for uptake and integration; and
- Step 3 - community engagement and recommendations for uptake and integration within the health system.

Additionally, capacity development through linking research to policy and practice for increased commitment and support by government to fund and implement further scale-up of this study in trauma facilities nationally will be threaded through the project.

### Study population and sampling procedure

The Applying Research to Policy and Practice for Health (ARCH) stakeholder mapping tool will be used to guide our mapping process [22] to identify stakeholders to participate in FGDs in WP1. FGDs will consist of approximately six to eight participants each and we anticipate holding between four and five FGDs or until theoretical saturation is reached. We will start by making a list of stakeholders in the trauma/injury prevention and alcohol policy fields according to four categories: academic, clinical, operational and policy stakeholders. Snowballing techniques from the initial core group will be used to identify additional stakeholders. Stakeholders will then be placed in a power-interest matrix based on the information outlined in the mapping [22] in order to categorize South African trauma and injury stakeholders according to their role in the professional landscape, to understand methods of engagement, and to lay out the proposed engagement strategy for the stakeholders throughout the project. FGDs will be exploratory and use a descriptive and contextual design [23]. Participants identified through this process will be invited to take part and be required to provide an informed consent before participation. FGDs with participants will take place over a two-month period and will take place virtually to accommodate stakeholders in different locations. These FGDs will be guided by a semi-structured interview sheet and run for between 45 and 60 minutes. They will be conducted by a trained facilitator in English.

For inclusion in the WP1 validation study consenting patients presenting at GSH will be 18 years and older; injured < six to eight hours prior to arrival at the facility. All new consecutive admissions meeting the inclusion criteria will be tested for alcohol use. This will include referrals from primary and district hospitals for specialized care at GSH. Unconscious, ventilated patients, and intoxicated patients with a breathalyzer (BrAC) test result > 0.10mg/l who will be regarded as too intoxicated for informed consent, will require delayed consent to participate. Patients with severe cognitive impairment will be excluded. If capacity to consent later is not regained, patients will be excluded. The required sample size to validate and assess the diagnostics’ performance is estimated at 1000 patients. This is based on the eligibility criteria and a targeted sample of cases stratified into a five-level category variable of intoxication and ICD-10 Y91 severity (Supp 1). From prior studies [24], we expect that 40% will be ineligible for study entry and a further 60% of the 600 eligible participants will have no alcohol detected or below 0.05g/100ml as the legal driving limit. The remaining four groups with a positive alcohol detection (“a patient who has a BAC reading of 0.05g/100ml” or above) will be closely monitored to ascertain that minimum of 60 cases per BAC vs ICD-10 Y91/diagnostic measurements code category is captured. Using the expected 60%/40% split in zero vs positive for alcohol we expect 360 zero alcohol and 240 alcohol-related cases, we do however expect that the distribution will vary across BAC categories.

Based on these assumptions, and to test a strict margin that the null hypothesis (k_0_=0.7) and the alternative hypothesis (k_1_=0.8) will be considered as substantial agreement, further input variables to determining the sample size included the 5 categories with frequencies equal to 0.6 (proportion of zero alcohol cases); and 0.1 (for each proportion of the 4 remaining alcohol positive categories). Based on this we are expecting a sample size of 396 participants at 90% power. This power calculation is based on a significance level of p<0.05. Hence, we expect our targeted sample of 600 eligible cases to be enough.

The same sampling strategy will be applied to select patients for the WP2 field testing of validated alcohol diagnostic tools in a district hospital trauma setting and estimated at approximately 1000 patients. The alcohol diagnostic tools will be used in separate data collection periods, for ease of utilization. As the selected district hospital has a monthly average caseload of approximately 561 cases [18], we will plan to collect an approximate sample of 400 eligible cases for one alcohol diagnostic tool (breathalyzer), followed by a week’s break, then for an additional 400 eligible cases to test a second diagnostic tool (Alcostick™) over the study’s planned duration of 90 days.

For WP3 hospital staff from the validation of alcohol diagnostics and field testing will be invited to participate in FGDs (Step 1). These will follow the same procedures described previously for the prioritization of methods and we will include a summary of relevant updated study information. In Step 2, national/provincial policy and health management stakeholders and the hospital staff from Step 1 will be invited to participate in a feedback workshop to lobby for uptake and implementation nationally and provincially. They will be complemented by approximately 30 key public health, clinician, health management, injury prevention and policy stakeholders, identified using the ARCH stakeholder mapping tool. Step 2 will be held in a hybrid virtual and in person workshop at the SAMRC in Cape Town after the FGD data has been analysed. For Step 3, we will work with SAMRC Corporate and Marketing Communications division for policy brief, infographic and short video design/production. This is for dissemination and engagement with community stakeholders, for research translation and recommendations to national/provincial policy and health management stakeholders, suggesting uptake and integration to other hospital trauma facilities.

### Data collection

The FGDs in WP1 will be conducted to explore stakeholder’s knowledge and views on current alcohol indicators collected in trauma settings and gaps in the collection of indicators. We will also obtain opinions from these stakeholders about diagnostic tools, implementation barriers, facilitators, feasibility, acceptability and the appropriateness of collecting routine and reliable injury surveillance and alcohol-related harm. Findings from this sub-study will inform the study on validation of alcohol diagnostic measures.

For the validation of alcohol diagnostics at GSH (WP 1), a draft survey questionnaire was adapted from the WHO Collaborative study on Injuries and Alcohol, a study validated across multi-country sites globally[24]. Information on the injury intent, mechanism, clinical screening according to ICD-10 code Y91, and breathalyzer analysis were retained with additions including the two alcohol diagnostic tools (the withdrawal of blood for testing and the Alcostick™ finger prick measure), time of blood withdrawal and finger prick, the South African Triage Scale^1^ (SATS), an indication of delayed consent, referring hospital name (if applicable) and hospital folder number.

Fieldworker study nurses will be employed and trained on informed consent and completion of the survey content, and the questionnaire/data capturing on Kobotools platform [27], using the study’s electronic Tablet devices. Fieldwork will occur over a 3-month period, with regular monitoring of the caseload to inform any adjustments to the schedule, until the targeted sample of 1000 cases are achieved, of which 600 cases are estimated to be eligible. Study nurses will be required to work between 8am and 4pm during a work week and between 5am and 7pm on weekends when required.

Blood samples will be couriered on a regular basis to a Pathcare laboratory near GSH. Preservation of the alcohol will be done by centrifuging the samples within 2 hours of blood withdrawal, to separate the plasma from the serum and to have it sealed in a separately labelled tube for courier and analysis at a second Pathcare facility identified for blood alcohol testing.

For WP2 the survey questionnaire will be revised for the field testing phase of validated alcohol diagnostics with the addition of questions on the body region injured (head, face, neck, thorax, etc.), the nature of injury (fracture, cut, bruise, concussion, etc.), the selected alcohol diagnostic tools (see Table 1), the patient’s drinking history prior to injury, a self-assessment on alcohol intoxication, and information on socio-economic status.

**Table 1.** Summary of alcohol diagnostic screening tool measures.

| Alcohol diagnostics | Data source |
| --- | --- |
| **Clinical assessment:** An observational assessment of alcohol intoxication conducted by a trained nurse or medical doctor. The assessment measures severity of speech impairment, motor coordination, behavioural disturbances, etc. through use of a Likert scale (none – very severe). | WHO Evidence of alcohol involvement determined by level of intoxication (ICD-10 Y91 codes)[25] |
| **Active breathalyzer testing:** The blood alcohol concentration measured in Breath Alcohol (BrAC) mg/l in exhaled breath through a straw. The minimum breath alcohol content detected is 0.03mg/l BrAC. Alcohol can be detected up to 6-12 hours since last drink.<br>**Passive breathalyzer testing:** Exhaled breath for ventilated patients to indicate the presence or absence of breath alcohol (Y/N) | Dräger: SANAS accredited breathalyzer screener used by blowing through a sterile mouthpiece for a digital reading (**Active**)<br>**Passive:** indicate the presence or absence of breath alcohol (Y/N) |
| **Surescreen Alcostick™:** Accurate measurement of capillary whole blood samples collect via finger prick, using a lancet. Results displayed in %BAC, mg/L or µg/dL BrAC. Lowest level of detection is 0.03% BAC. | Surescreen Alcostick™: rapid blood alcohol meter [19] |
| **Blood sample :** Detecting the presence or absence of ethanol in the blood [26]. Venous blood collected in a Sodium Fluoride tube; alcohol levels reported in g/dL. | Testing with venous blood sample. |

<sup>1</sup> The South African Triage Scale (SATS) can be assessed by doctors, registered nurses, and enrolled nurses/nursing assistants. The SATS has four priority levels (with corresponding targeted treatment times): Red (immediate treatment), Orange (treat in <10 min), Yellow (<1hour), Green (<4hours), and death indicate by a 5<sup>th</sup> category: Blue (death certification by a doctor in <2hours) (Western Cape Government, 2012).

Fieldwork will be based on an idealized week, with a selection of hours across day- and nighttime, during the 3-month period. This is to ensure that the data is representative in terms of the facilities’ operating conditions, which is expected to change by time of the month, hour, week- or weekend day, etc. to assess the suitability of the alcohol diagnostic tools for possible national scale-up.

In WP3 FGDs with participants in Step 1 will take place virtually to accommodate stakeholders in different locations. Interview guides will consist of open-ended questions to explore participants’ views on the recommended methods, their thoughts on the requirements for implementation (provincially and nationally) and what the role of the measure would be in health care provision more broadly. Specific areas to discuss will include 1) intervention content, 2) intervention delivery, 3) strategies for addressing possible barriers to intervention delivery, 4) research questions around acceptability, feasibility and sustainability of measuring alcohol-related trauma, and 5) strategies to generate local stakeholder buy-in. These areas of discussion will ensure appropriate information is collected that will inform national stakeholders on the benefits for adoption of the recommended alcohol diagnostic screening tools for larger implementation [28].

For Step 2, findings from WP1 and WP2, as well as the FGD results from WP3 (Step 1) will be shared in a workshop with researchers; clinicians and hospital managers; traffic officials; emergency medical services; police, pathology and rescue services; and national and provincial policy makers. The output will be a framework to guide implementation and scale-up of routine diagnostic measuring of alcohol-related trauma. Workshop participants will be asked to evaluate the usefulness of the workshop and their satisfaction with the outcome.

### Data Analysis

WP1 and WP 3 FGDs will be audio recorded and transcribed verbatim. Thematic analysis will be conducted based on deductive themes focusing on the exploration of current practices, implementation barriers, facilitators, feasibility, acceptability and appropriateness of conducting alcohol diagnostics in public health facilities (WP1.1) and on acceptability, feasibility and appropriateness of the recommended measure (WP 3). The data will be managed using qualitative data analysis software NVivo, V.12. and will be presented in line with COnsolidated criteria for REporting Qualitative research (COREQ) guidance for reporting qualitative research [29].

For the validation of alcohol diagnostics in WP1 and for WP2, data collection will be regularly monitored and managed by the study statistician. The blood alcohol analysis results will be merged to the main database using the unique case ID. The correlation coefficients of the continuous alcohol diagnostic measurements and the blood alcohol tests will be reported and represented graphically. A Kappa statistic [30] will be used to assess the level of agreement^2^ between BAC categories and the various alcohol diagnostic tests and the clinical assessment as per ICD-10 Y91 codes. Taking into consideration that the alcohol diagnostic measures are ordinal responses, we will utilize ordinal regression to model the Receiver Operating Characteristic (ROC) curve [31, 32]. This diagnostic test plots sensitivity against specificity of the alcohol diagnostics measures against the gold standard BAC. The area under the ROC curve will also be used to characterize the accuracy of the diagnostic tests, providing all information on its performance, instead of only a single estimate of the test’s sensitivity and specificity. The trade-offs between the ROC curve’s sensitivity and specificity can then be assessed to inform a decision threshold [33, 34] for the alcohol diagnostic methods to be used in phase 2. Any level above 70% sensitivity and specificity will be acceptable. If multiple tests meet the same criteria, we will use the area under the curve to determine the best test. We will however proceed to test the feasibility of use for both the breathalyzer and the Alcostick™ if they meet the specified sensitivity and specificity criteria. Besides reporting the results of validating the alcohol diagnostic tools, descriptive statistics for analysis will include age and gender, followed by analysis on severity of the injury (assessed by the triage scale), intent of the injury (violence, road traffic, unintentional and self-harm), the related mechanism of injury (i.e. gunshot, stabbing, pedestrian, driver, fall, etc.) and BAC categories. Data analysis will be conducted on STATA version 17 (Stata/IC 17, 2021). Data will be presented in line with STROBE guidance for reporting cross-sectional studies [35]

For the field testing (WP 2), we will follow the same data management and analysis procedures and packages as for the validation of alcohol diagnostics. Further analysis will include patient demographics, the nature of injury (fracture, sprain, open wound, burn, etc.) and body region injured, the intent of the injury (violence, road traffic, unintentional and self-harm) and the related mechanism of injury (i.e. gunshot, stabbing, pedestrian, driver, fall, etc.) and triage scale. We will use ordinal regression to analyze the severity of injuries (as indicated by the triage scale) as the outcome, and level of intoxication (indicated as 0.05g/100ml and above) as one of the independent variables. In addition, a multinomial regression will be used to assess the association between the outcome as type of injury with the level of alcohol intoxication. As information on drinking prior to injury will be recorded to determine the type of alcohol and volume consumed, the dose-response relation between the number of drinks consumed within 6 hours leading up to the injury and the relative risk (RR) of being injured will be analysed and reported. Socio-economic information on employment status, household income and suburb of residence will be captured and used to inform alcohol policies.

For the workshop evaluation form in WP 3, we will sum each evaluation item to create scores for the evaluation form and will summarize using mean with the standard deviation or median with interquartile range.

### Patient and public involvement

Clinicians are an important participant group and were involved in the initial design of the study and as co-investigators. We will be using a participatory approach through stakeholder engagement initiated in the prioritization of methods to include community and patient representation; then incorporating a synthesis of findings from the validation of alcohol diagnostics and suitability testing to conduct qualitative research on the utility of the recommended alcohol diagnostic method; and where feedback will be sought on requirements for uptake and integration within the health system which has a specific focus on community-based participatory research for study synthesis and recommendations. This final uptake activity is iterative and dependent on the outcomes of the formative work. Therefore, the study overall is geared to soliciting participant codesign.

## OUTCOMES

A validated method(s) of diagnosing alcohol-related injury and violence for use in clinical settings.

## SUMMARY AND DISCUSSION

There is a lack of routine and reliable injury surveillance and specifically alcohol-related harm data in South Africa to respond to the related trauma burden. Additionally, research to develop systems or reliable mechanisms to test alcohol-relatedness of trauma and to monitor the impact of interventions is lacking. Alcohol is an established risk factor for violence and injuries and accurately monitoring alcohol-relatedness in response to interventions, policy changes, etc. will be necessary to evaluate effectiveness -as suggested by the WHO SAFER strategy of “*Monitoring*”, one of three essential strategies aimed at government officials for the purpose of developing evidenced-based alcohol policies and action plans to address alcohol harm [36]. The prominent role of alcohol in the trauma setting has become particularly pronounced during the COVID-19 lockdown, and this study will provide crucial evidence needed for effective measurement of alcohol-related trauma to improve injury surveillance and clinical management. Through this study we hope to identify and validate the most appropriate method(s) of diagnosing alcohol-related injury and violence in clinical settings. Findings from this study are likely to be highly relevant and could influence our primary beneficiaries – policy makers and senior health clinicians – to adopt new practices and policies around alcohol testing in injured patients.

## ETHICS AND DISSEMINATION

Ethical approval for the study has been granted by the Research Ethics Committee of the South African Medical Research Council (EC005-2/2022) and approval from the Western Cape Health Department. Findings will be disseminated to relevant national and provincial government departments, policy experts and clinicians. We will publish findings in scientific journals and engage in media advocacy and share findings with our stakeholders, including community representatives, non-profit partners and civil society organisations.

## Data Availability

All data produced in the present study are available upon reasonable request to the authors

## Authors’ contributions

PPW and MRP conceptualized the paper. PPW, RM and MMP developed the first draft. All authors edited the subsequent versions of the draft. All authors have reviewed and accepted the final version of the protocol and given their permission for publication.

## Footnotes

1 The South African Triage Scale (SATS) can be assessed by doctors, registered nurses, and enrolled nurses/nursing assistants. The SATS has four priority levels (with corresponding targeted treatment times): Red (immediate treatment), Orange (treat in <10 min), Yellow (<1hour), Green (<4hours), and death indicate by a 5^th^ category: Blue (death certification by a doctor in <2hours) (Western Cape Government, 2012).

2 0.01–0.20 represents zero to slight agreement, 0.21–0.40 represents fair agreement, 0.41– 0.60 as moderate, 0.61-0.80 is considered substantial agreement (Cohen, 1968).

