## Supplemental 1 for "A multi-method study to identify and validate alcohol diagnostics for injury-related trauma in South Africa: a study protocol"

Supp 1

**Targeted sampling strategy with minimum sample requirement, prior to eligibility assumptions by BAC reading and Y91 code**

| | **BAC reading (g/100ml*) by severity of alcohol intoxication** | | | | | |
|---|---|---|---|---|---|---|
| | None[1] (0.000-0.049) | Mild (0.050-0.099) | Moderate (0.100-0.199) | Severe (0.200-0.299) | Very severe (0.300 +) | Min. Total |
| Zero alcohol | 360 | | | | | 360 |
| Y91.0 Mild alcohol intoxication | | 60 | | | | 60 |
| Y91.1 Moderate alcohol intoxication | | | 60 | | | 60 |
| Y91.2 Severe alcohol intoxication | | | | 60 | | 60 |
| Y91.3 Very severe alcohol intoxication | | | | | 60 | 60 |
| Min. Total | | | | | | 600 |

***BAC readings for BrAC collected by the AlcostickTM and Breathalyzer will be converted to g/100ml during the analysis stage.

[1] A person is assumed to be under the influence of alcohol when the quantity consumed exceeds the alcohol tolerance of an individual, and causes impairment in mental and physical ability. These effects are usually influenced by several factors such as metabolism, consumption history, body fat content and pre-existing medical conditions (Greeley & McDonald, 1992). Hence, the corresponding scale is used for Supp 1 and BAC levels within the legal South African driving limit of <0.05g/100ml will be excluded for the analysis of "alcohol intoxication".
